# Bridging Language Gaps in Neurology Patient Education Through Large Language Models: a Comparative Analysis of ChatGPT, Gemini, and Claude

**DOI:** 10.1101/2024.09.23.24314229

**Authors:** Mahdi Haq, Muhammad Mushhood Ur Rehman, Mohamed Derhab, Reeda Saeed, Junaid Kalia

## Abstract

This study evaluates the capability to translate neurology patient education material using three Large Language Models (LLMs) - ChatGPT-4 Omni, Gemini 1.5 Pro, and Claude 3.5 Sonnet. Five neurological conditions (Bell’s palsy, multiple sclerosis, stroke, migraine, and epilepsy) were translated from English into Spanish, Urdu, and Arabic. The translations were assessed by physicians using four metrics: accuracy, clarity, comprehensiveness, and readability at a 6th grade level. Results showed that Claude outperformed both ChatGPT and Gemini overall, particularly excelling in Spanish and Urdu translations, while Gemini led in Arabic. All LLMs demonstrated superior performance in Spanish compared to Urdu and Arabic. This study highlights the potential of LLMs in enhancing patient education across languages, while also identifying areas for improvement in translation accuracy and readability.

## Introduction

Large Language Models (LLMs) have taken the world by storm since the release of OpenAI’s ChatGPT in November 2022. Since then, there have been numerous studies on the utility and application of LLMs in healthcare. From generating patient education material, to using artificial intelligence to chat with patients in real time, to developing algorithms for reading radiology scans, AI has been at the forefront of new initiatives in healthcare. The ability of LLMs to understand and generate text has opened the doors of immense potential in healthcare. However, the introduction of LLMs in healthcare raise concerns as well, especially regarding accuracy, safety, and privacy. This study aims to look at the capability of LLMs to translate patient education material that is accurate, safe, and effective. This can translate over to improving patient outcomes. LLMs are even able to answer examination level medical questions with ease^1^. Importance of patient education for health outcomes, significant clinical benefits have been seen as a result of patient education in diseases such as diabetes, heart disease, and rheumatoid arthritis^2^.

## Objectives

The objective of this study is to evaluate and compare the ability of three different LLMs to translate neurology patient education material from English into three different languages: Spanish, Urdu, and Arabic. The three LLMs being tested are ChatGPT-4 Omni by OpenAI, Gemini 1.5 Pro by Google, and Claude 3.5 Sonnet by Anthropic.

## Methodology

This is a cross sectional study. We used pre-existing patient education material in English from a neurological research institute, NeuroCareAI. We chose patient education material for five common neurological conditions to put to the test: Bell’s palsy, multiple sclerosis, stroke, migraine, and epilepsy. Each of the LLMs translated all five diseases into three languages: Spanish, Urdu, and English. The prompt we used for each LLM was the same: ‘Translate this entire document into Spanish/Urdu/Arabic’. Once the translation was complete, each translation was evaluated by three physicians who were native speakers of that respective language. Four metrics were used in the evaluation: accuracy, clarity, comprehensiveness, and readability at a 6th grade level. Each metric was graded on a Likert scale from 1 to 5.

### Likert scale used in all evaluations

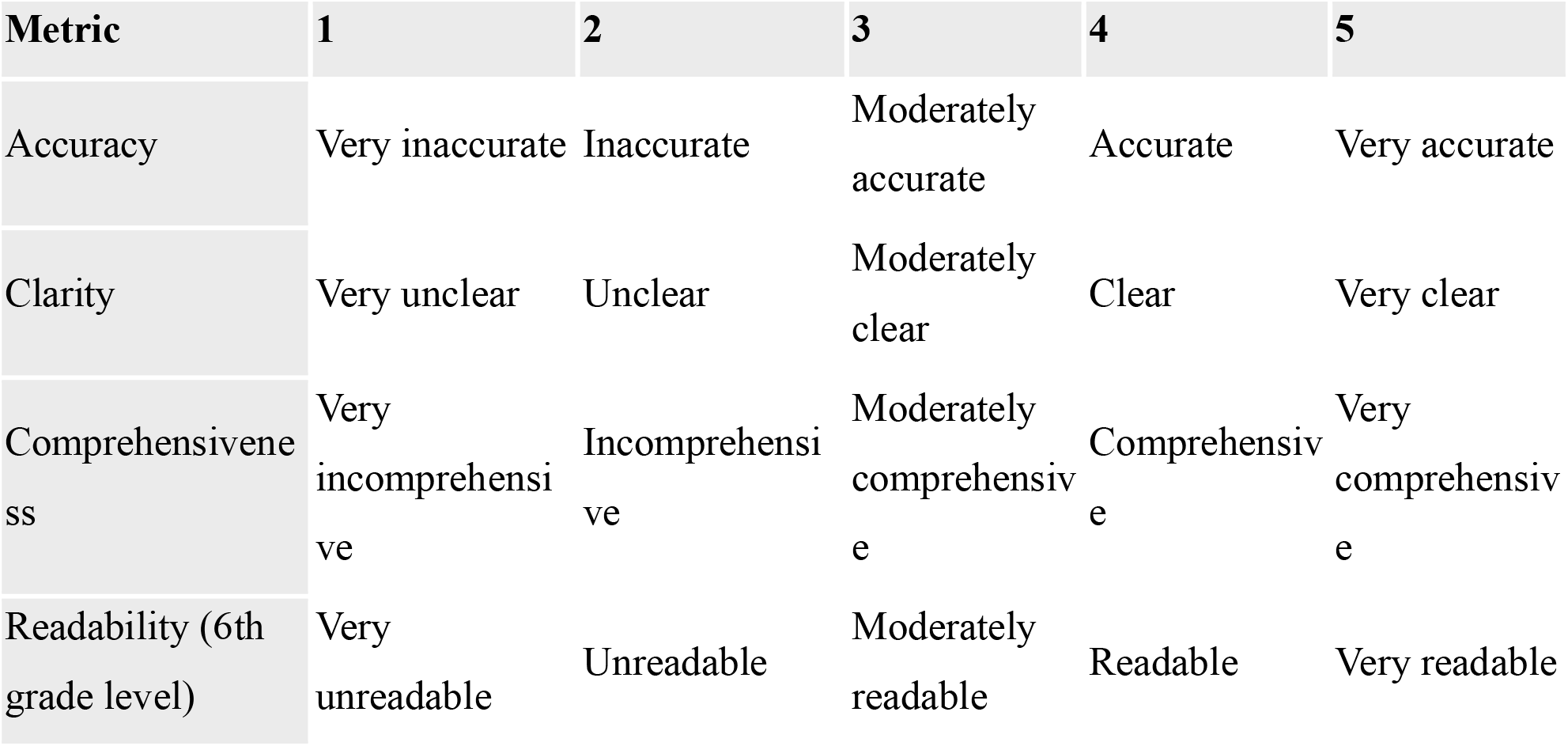

## Results

Claude outperformed ChatGPT and Gemini overall, with a total score of 737 out of 900, with Gemini getting 694 out of 900, and ChatGPT with the least score of 669 out of 900. Claude was the frontrunner for the Spanish and Urdu translations, but had a lower score than Gemini for the Arabic translations. When looking at accuracy, Claude was the most accurate for Spanish and Arabic, but ChatGPT performed better for the Urdu translations. Regarding clarity, Claude was the best for Spanish and Urdu, but was outperformed by Gemini for Arabic. In the comprehensiveness metric, Claude once again performed the best for Spanish and Urdu, and tied with Gemini for Arabic. Lastly, when it came to readability, Claude was the best for Spanish, tying with ChatGPT for Urdu, and Gemini taking the lead for Arabic. Interestingly, all three LLMs performed much better for the Spanish translations overall, compared to the Urdu and Arabic translations.

**Figure 1.**
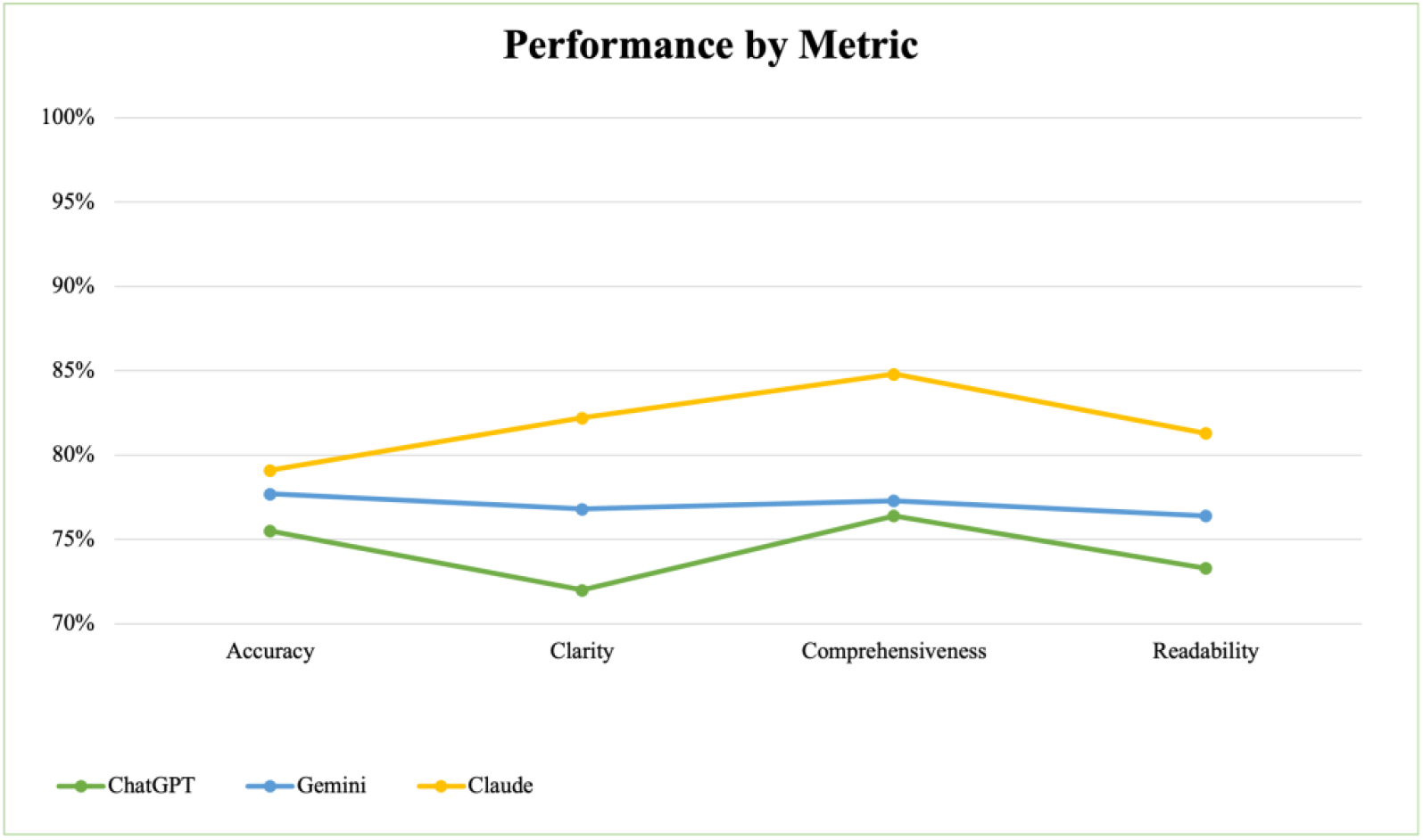

**Figure 2.**
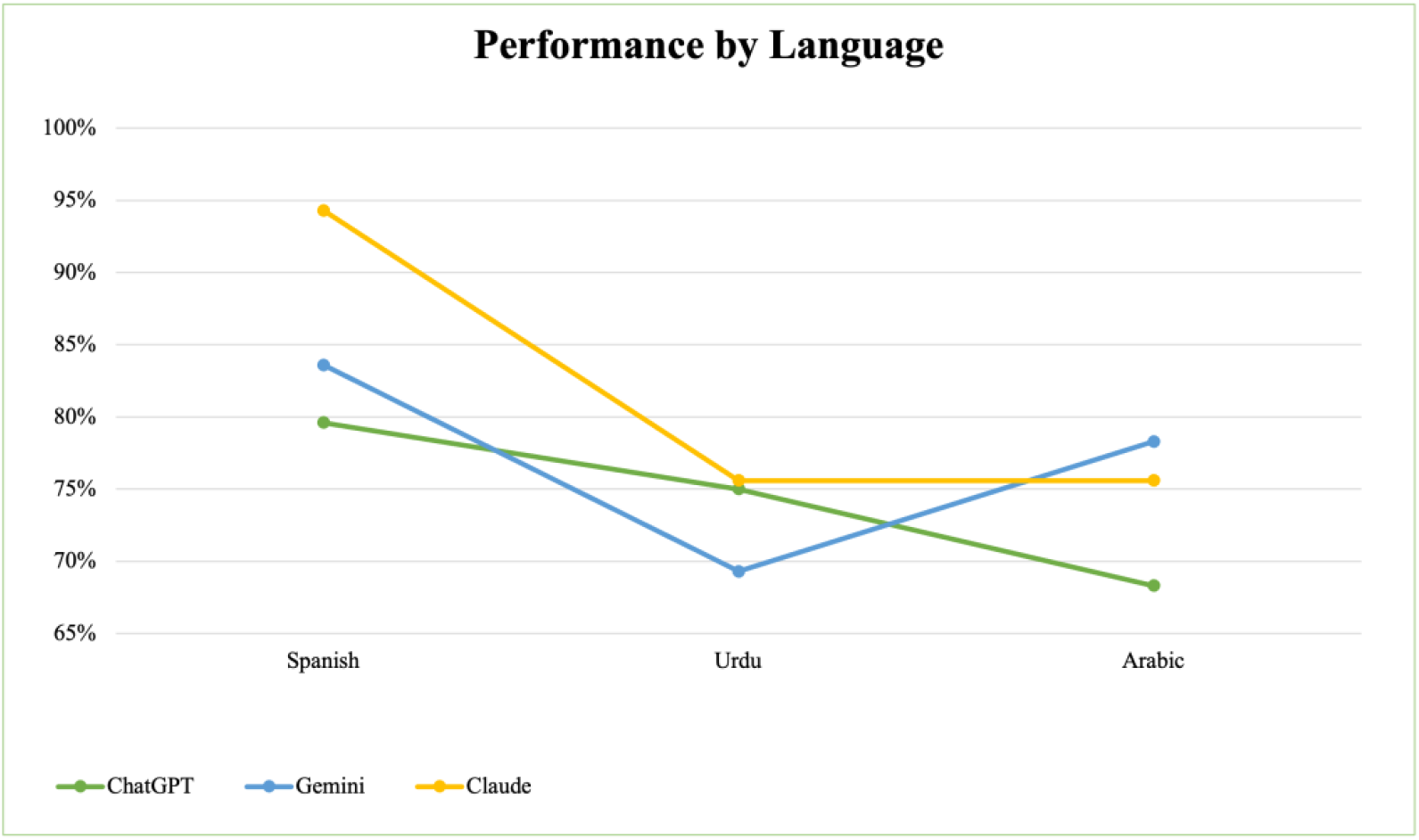

## Discussion

The three LLMs that we tested are currently among the most popular ones. Anthropic’s Claude 3.5 Sonnet had the best performance overall in our study. All three LLMs also performed much better when it came to translating the patient education material into Spanish, as compared to Urdu and Arabic. This suggests that LLMs are better at translating patient education material from English to other similar languages. In a previous study regarding LLM generated patient education material, ChatGPT-4 performed the best when compared to Google’s Bard (now known as Gemini) and Microsoft’s Bing^3^. There are currently not many studies assessing the difference in capability of LLMs in accurately translating existing patient education material into different languages. In another study, ChatGPT-4 outperformed Google’s Bard in converting patient education material into a more readable version^4^.

The main limitation of our study was that we did not have an objective tool to analyze the readability of Urdu and Arabic. There are about ten tools that exist to objectively determine the readability of a text in the English language, however, no such tools exist or are publicly available for Urdu and Arabic. Another limitation was that we tested the translation capabilities for patient education material regarding only five neurological diseases, and not any other patient education material from other medical specialties. There is room for improvement in the LLMs capability to translate patient education material into Urdu and Arabic, while the existing capability for translating into Spanish is much more satisfactory. There is massive potential for LLMs to make quality patient education material available in different languages in which no quality patient education material exists. This can have a positive effect on health outcomes for those people that previously have not had patient education material available in their own language.

## Data Availability

All data produced in the present study are available upon reasonable request to the authors.

## Notes

### Competing Interest Statement

The authors have declared no competing interest.

### Funding Statement

This study did not receive any funding.

